# Worldwide association of lifestyle related factors and COVID-19 mortality

**DOI:** 10.1101/2021.08.14.21257136

**Authors:** Jingzhou Wang, Toshiro Sato, Atsushi Sakuraba

## Abstract

**Background:** Several lifestyle related factors such as obesity and diabetes have been identified as risk factors for Coronavirus disease 2019 (COVID-19) mortality. The objective of this study was to examine the global association between lifestyle related factors and COVID-19 mortality using data from each individual country.

**Methods:** The association between prevalence of seven lifestyle related factors (overweight, insufficient physical activity, smoking, type 2 diabetes, hypertension, hyperlipidemia, and age over 65) and COVID-19 mortality was assessed by linear and multivariable regression among 186 countries. The cumulative effect of lifestyle related factors on COVID-19 mortality was assessed by dividing countries into four categories according to the number of lifestyle related factors in the upper half range and comparing the mean mortality between groups.

**Results:** In linear regression, COVID-19 mortality was significantly associated with overweight, insufficient physical activity, hyperlipidemia, and age ≥65. In multivariable regression, overweight and age ≥65 demonstrated significant association with COVID-19 mortality (P = 0.0039, 0.0094). Countries with more risk factors demonstrated greater COVID-19 mortality (P for trend <0.001).

**Conclusion:** Lifestyle related factors, especially overweight and elderly population, were associated with increased COVID-19 mortality on a global scale. Global effort to reduce burden of lifestyle related factors along with protection and vaccination of these susceptible groups may help reduce COVID-19 mortality.

## Introduction

Coronavirus disease 2019 (COVID-19) has infected over 33 million individuals and claimed at least one million lives across the globe.[1] Declared as a pandemic by the World Health Organization (WHO), COVID-19 has penetrated most countries, although the infection and death rates vary substantially across different countries. In China, where COVID-19 initially started, the infection and death rates are 59 and 3 per one million population, respectively.[2] Most Asian countries have rates in the range similar to China.[2] In many European countries, these numbers are much greater with infection and death rates in the range of 3000-6000 and 300-800 per million, respectively.[2] Such variations in infection and death rates may be explained by differences in government policies on lockdown or rates of compliance of social distancing and mask wearing by citizens. However, difference in demographics and the prevalence of lifestyle related factors may also contribute to variations in COVID-19 mortality.

Before the start of COVID-19 pandemic, there had been a silent but significant pandemic happening in the background, the obesity pandemic. As a major risk factor for non-communicable diseases, obesity substantially increases the risks of developing type 2 diabetes mellitus, hypertension, myocardial infarction, stroke, dementia, fatty liver disease, osteoarthritis, obstructive sleep apnea and several cancers. [3] The prevalence of obesity has been increasing in every country in the past 50 years, particularly in developing countries in South Asia, the Caribbean and southern Latin America. [4] It has been estimated that at least 10% of adults globally have body mass index (BMI) ≥30 kg/m2. [5] Over 30% of adults are obese in the United States, the country with the highest number of COVID-19 mortality. The COVID-19 pandemic has revealed a connection between communicable and non-communicable diseases, in which people with chronic lifestyle related factors such as obesity, diabetes and hypertension, which are common risk factors for cardiovascular diseases and chronic kidney diseases, seem to be vulnerable to COVID-19 infection.[6, 7] Obese patients were 3 times more likely to develop severe COVID-19 disease compared to patients with normal BMI.[6] Increased risk of intensive care unit (ICU) admission have been reported in younger patients who were obese. Furthermore, elderly people were shown to be at higher risk of COVID-19 deaths. The case fatality rate was greater in patients age ≥60 as compared to <60 (6.4% vs 0.32%).[8] However, these risk factors reported above were largely based on single-center studies or individual country’s own experiences.

There has been a lack of studies on the association between lifestyle related factors and COVID-19 mortality on a global scale. The current study aimed to address this gap by investigating the worldwide association between lifestyle related disease burden and COVID-19 mortality.

## Methods

We included seven lifestyle related factors in our present study. The data of global prevalence of hypertension, hyperlipidemia, smoking, overweight, and insufficient physical activity, were obtained from the 2015/2016 WHO handbooks.[9] The data on population percentage with age ≥65 was extracted from 2019 World bank.[10] Data of diabetes prevalence was obtained from the 2019 International Diabetes Federation.[11] Data of mortality (deaths per million population) due to COVID-19 was obtained from Worldometer, which is an international live world statistics working group that has been widely used in research.[2, 12, 13] All databases were searched on August 20, 2020. There were 186 countries with data of mortality and at least one other variable.

## Study aims and statistical analysis

The primary aim was to study the association between the seven lifestyle related factors and deaths due to COVID-19 by linear regression. Multiplicity adjusted P values following Bonferroni multiple comparison testing was performed. A corrected P-value of <0.05 was considered statistically significant. Multivariable regression was performed by applying backward elimination with a threshold of 0.05 for selection of most predictive variable(s). Differences were considered to be statistically significant when the P-value was <0.05.

The secondary aim was to assess the cumulative effect of lifestyle related factors on COVID-19 mortality. Countries were divided into 4 categories, having 0-1, 2-3, 4-5, or 6-7 risk factors according to the number of lifestyle related factors in the upper half range of the global ranking. The mean mortality among groups were compared by one-way analysis of variance. The trend in increasing mortality with greater cumulative risk factors was analyzed by Jonckheere-Terpstra test.

Data were analyzed by EZR (Saitama Medical Center, Jichi Medical University, Saitama, Japan),[14] which is a graphical user interface for R (The R Foundation for Statistical Computing, version 2.13.0, Vienna, Austria), and GraphPad Prism version 6 (San Diego, CA).

## Results

A total of 186 countries were available for analysis. Countries from all continents were included. There were 55, 45, 45, 28, and 13 countries from Africa, Asia/Oceania, Europe, North/Central America, and South America, respectively.

### Linear regression between lifestyle related factors and COVID-19 mortality

We first undertook linear regression analysis to study the association between each variable of interest and number of deaths due to COVID-19 per million population (**Table 1**). COVID-19 mortality showed significant positive association with the proportions of overweight (R^2^ = 0.15, P <0.001), age ≥65 (R^2^ = 0.11, P <0.001), insufficient physical activity (R^2^ =0.04, P = 0.0077), and hyperlipidemia (R^2^ = 0.16, P <0.001). No statistically significant association was found between COVID-19 mortality and the prevalence of diabetes, hypertension or smoking in linear regression. The results of the linear regressions of proportion of overweight and age ≥65 with COVID-19 mortality are shown in **Figure 1A, B**.

**Table 1.** Summary of linear regression and multivariate analysis between each variable and mortality due to COVID-19

|  | Linear regression |  |  | Multivariable regression with stepwise selection |
| --- | --- | --- | --- | --- |
|  | Adjusted R<sup>2</sup> | Regression | P value | P value |
| **Hyperlipidemia** | 0.16 | 4.80 | 0.000000070 |  |
| **Type 2 diabetes** | -0.0042 | -2.04 | 0.58 |  |
| **Hypertension** | 0.0032 | -3.35 | 0.22 | 0.0065 |
| **Insufficient physical activity** | 0.044 | 3.50 | 0.0077 |  |
| **Overweight** | 0.15 | 4.25 | 0.00000017 | 0.0039 |
| **Smoking** | -0.00050 | -2.63 | 0.33 |  |
| **Age ≥65 years** | 0.11 | 9.23 | 0.0000034 | 0.0094 |

**Figure 1.**
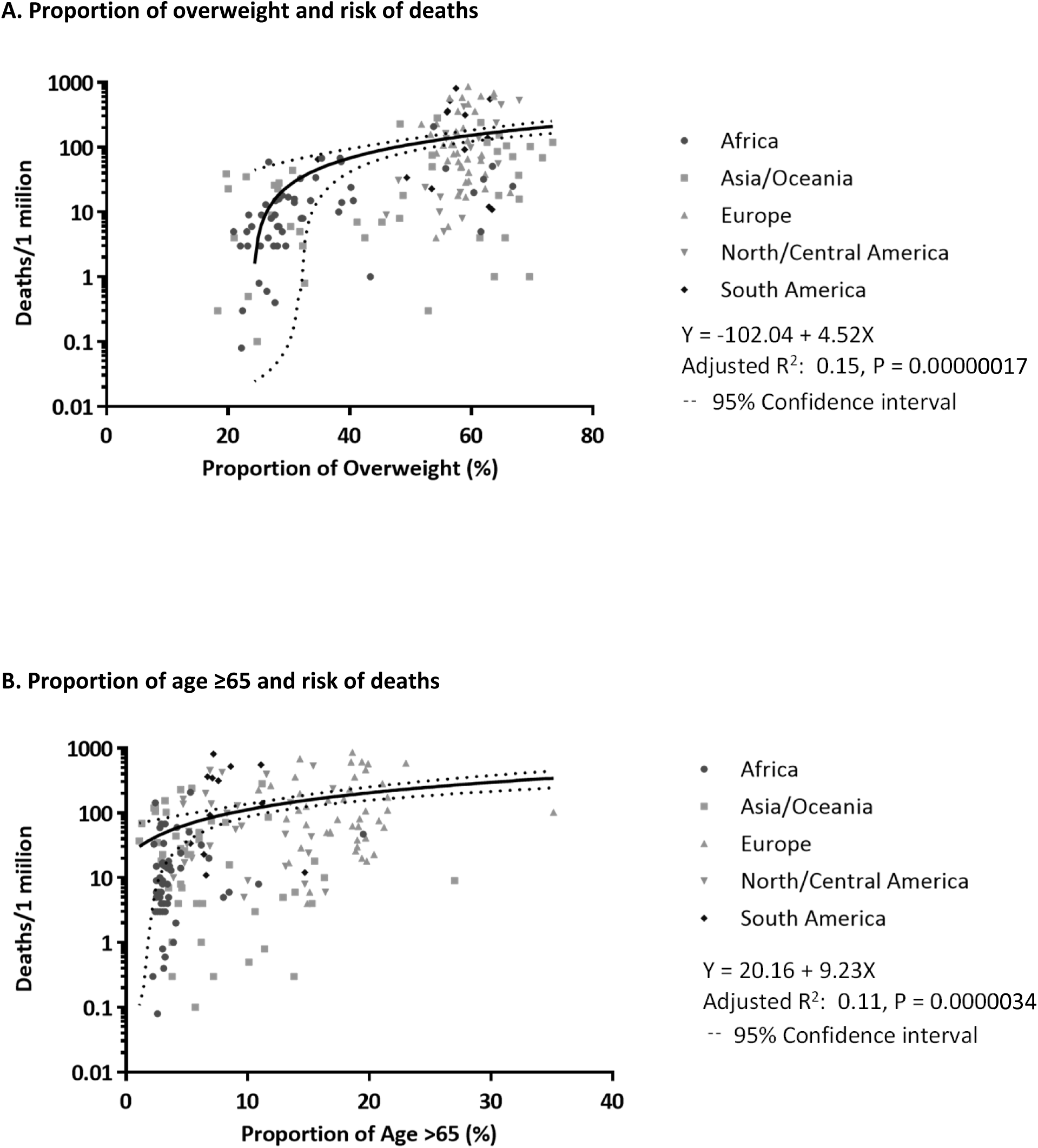
Linear regression between the proportion of overweight and age >65 and mortality due to COVID-19. **A.** The proportion of population of overweight amongst countries strongly correlated with the number of deaths per 1 million population (adjusted R^2^ 0.15, P = 0.00000017). **B.** The proportion of population age ≥65 amongst countries correlated with the number of deaths per 1 million population (adjusted R^2^ 0.11, P = 0.0000034).

### Multivariable regression between lifestyle related factors and COVID-19 mortality

We next performed multivariable regression with stepwise selection to estimate the relationship between these seven lifestyle related factors and COVID-19 mortality. The proportion of overweight and age ≥65 demonstrated significant positive association with COVID-19 mortality (**Table 2**, P = 0.0039 and 0.0094, respectively) on multivariable regression. On the other hand, the prevalence of hypertension showed a weak negative association with COVID-19 mortality (P = 0.0065).

### Cumulative influence of lifestyle related factors on COVID-19 mortality

We next studied the cumulative effect of lifestyle related factors on the risk of COVID-19 mortality. Countries were divided into four categories according to the numbers of lifestyle related factors in the upper half range of the global ranking (0-1, 2-3, 4-5, or 6-7 risk factors). As shown in **Figure 2**, countries with more risk factors demonstrated greater mortality due to COVID-19 (P = 0.015 between 0-1 vs. 4-5, Jonckheere-Terpstra test for trend P <0.001) suggesting that there might be synergistic effects of these risk factors. Interestingly, the difference between countries carrying 4-5 or 6-7 risk factors was small.

**Figure 2.**
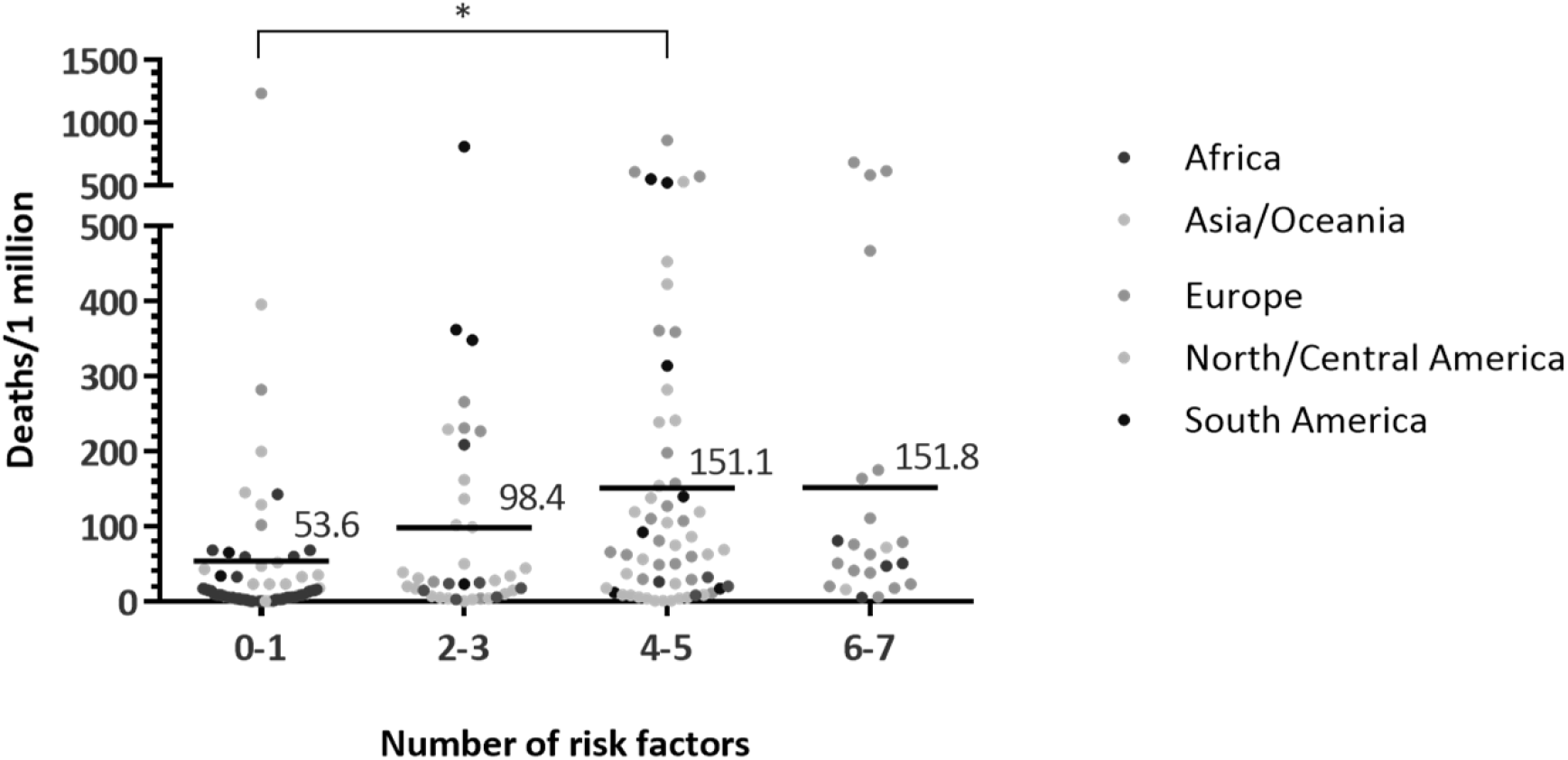
Cumulative number of lifestyle related risk factors and risk of deaths. Countries with more risk factors demonstrated greater mortality due to COVID-19 (P = 0.015 between 0-1 vs. 4-5, Jonckheere-Terpstra test for trend P <0.001).

## Discussion

The present study shows that the prevalence of overweight and elderly population were significantly associated with COVID-19 mortality at a global level. Furthermore, countries with higher burden of lifestyle related factors suffered a greater casualty due to COVID-19. The burden of lifestyle related factors reflects the life time cumulative effects of unhealthy lifestyles, which increases the risk of developing cardiovascular diseases, strokes, and certain types of cancer.[15] These risks may persist over the life span of adulthood, but have recently been reported in younger populations.[16] Lifestyle related factors impose a considerable economic burden on health services. The results of our study highlight that lifestyle related factors may also influence individual country’s mortality against a communicable infection caused by a global pandemic.

Significant variation in cases of infections and deaths due to COVID19 among countries have been reported.[2] For instance, countries in North/South America and Europe have shown high mortality rates ranging from 300-800 per million population. On the other hand, Asian countries such as South Korea, Japan, and Thailand have shown much lower mortality rates ranging from 0.8-9 per million population. While such variations are partially explained by differences in government policies, and citizen’s compliance on social distancing and mask wearing, the results of our study suggest that each country’s demographics and burden of lifestyle related factors and co-morbidities also play a role. This suggests that severe COVID-19 shares many of the risk factors with atherosclerosis and cardiovascular diseases, and prevention of chronic lifestyle related factors by healthy lifestyles and population health approach may help mitigate casualty caused by COVID-19.

The present study supports the association between obesity and COVID-19 mortality. Adipose tissue increases the production of inflammatory cytokines, such as IL-6, in which its blockade has demonstrated promising results as a COVID-19 therapeutic option.[17] Obesity increases the risk of diabetes and hypertension, which upregulate the expression of ACE2 protein that COVID-19 binds when entering cells.[1] The lock-down to mitigate the spread of COVID-19 has led to quarantine weight gain and poor control of diabetes,[18] so urgent action to prevent obesity is needed to reduce further negative influence during the subsequent waves of pandemic.

The association between older age and COVID-19 mortality is not surprising. Influenza is disproportionately more lethal in elderly persons[19] and COVID-19 has also proven devastating in the geriatric population. Not only do elderly people have weakened immune system, many also live in nursing homes, where high resident density further contribute to the COVID-19 spread.[20] Elderly nursing home residents accounted for approximately 25% of the documented deaths due to COVID-19 in many countries.[21]

Although insufficient physical activity and hyperlipidemia were associated with COVID-19 mortality in linear regression, the associations were not statistically significant in multivariable analysis. Similarly, we did not identify any significant association between COVID-19 mortality and the prevalence of diabetes or hypertension suggesting that each lifestyle related factor has different degree of influence on COVID-19 risk.[22] The lack of association may be caused by differences in other non-demographic factors, such as social distancing and mask wearing, across countries that contribute to COVID-19 mortality. However, when analyzed by cumulative burden of lifestyle related factors, countries with greater numbers of these factors had more casualty due to COVID-19. Chronic lifestyle related factors have been causing economic burden by increasing morbidity and mortality of atherosclerosis, cardiovascular diseases, and cancers,[23] but the results of our study suggest that it may also influence the outcome of viral pandemics. Global and governmental leaders need to urgently work on improving the behavior and lifestyle of the citizens and promote healthy lifestyles because the pandemic is far from over and may last for a few years. Furthermore, a similar degree of casualty is expected to occur if we encounter a future pandemic without reduction of the burden of lifestyle related diseases.

The present study has several limitations. The use of different datasets for variables of interest may result in heterogeneous measurements due to variations in data collections, so the reported outcomes may not reflect the true prevalence in real time. Second, global investigation of COVID-19 mortality is subjected to differences in testing rate across different countries, in which underdeveloped countries may under report the true infection rates due to limitation of testing capacities. This is why the current paper chose death per million population as the main outcome, instead of case fatality rate or number of infections, as the former outcome depends less on testing rate.[12, 24] We are aware that our analysis is preliminary as the COVID-19 pandemic is still progressing and the infection may not have fully penetrated in some countries, so the mortality rate may change among countries. We plan to perform a follow-up analysis to see whether our results can be verified with the updated death toll after the pandemic has fully passed. Furthermore, COVID-19 associated death rates may be influenced by other factors such as differences in government policies on lockdown, rates of compliance of social distancing and mask wearing by citizens, or host genetic factors that control susceptibility to infections.[25] Lastly, we are aware that while our observational study demonstrated association, this does not mean causation exists between the variables and the outcome.[13] Further studies investigating causal relationship are necessary in this field.

In summary, we studied the association between lifestyle related factors and mortality due to COVID-19 and demonstrated that countries with higher prevalence of overweight and elderly population had increased COVID-19 mortality. Furthermore, countries with higher prevalence of various lifestyle related factors suffered a greater casualty due to COVID-19. Focused measures to protect and vaccinate these susceptible groups early may help reduce COVID-19 mortality. Global effort to reduce burden of lifestyle-related factors may be a promising strategy to reduce mortality from viral pandemics.

## Data Availability

Link to databases are provied in the manuscript.

https://data.worldbank.org/indicator/SP.POP.65UP.TO.ZS?most_recent_value_desc=false

https://www.diabetesatlas.org/data/en/indicators/2/

https://www.who.int/data/gho

## Figure legends

**Figure 1**. Linear regression between the proportion of overweight and age >65 and mortality due to COVID-19. **A**. The proportion of population of overweight amongst countries strongly correlated with the number of deaths per 1 million population (adjusted R^2^ 0.15, P = 0.00000017). **B**. The proportion of population age ≥65 amongst countries correlated with the number of deaths per 1 million population (adjusted R^2^ 0.11, P = 0.0000034).

**Figure 2**. Cumulative number of lifestyle related risk factors and risk of deaths due to COVID-19. Countries with more risk factors demonstrated greater mortality due to COVID-19 (P = 0.015 between 0-1 vs. 4-5, Jonckheere-Terpstra test for trend P <0.001).

